# Antimicrobial susceptibility testing reveals reduced susceptibility to azithromycin and other antibiotics in *Legionella pneumophila* serogroup 1 isolates from Portugal

**DOI:** 10.1101/2023.09.25.23296014

**Authors:** Corrado Minetti, Rachael Barton, Caitlin Farley, Owen Brad Spiller, Raquel Rodrigues, Paulo Gonçalves

## Abstract

**Background:** Although not fully investigated, studies show that *Legionella pneumophila* can develop antibiotic resistance. As there is limited data available for Portugal, we determined the antibiotic susceptibility profile of Portuguese *L. pneumophila* serogroup 1 (LpnSg1) isolates against antibiotics used in the clinical practice in Portugal.

**Methods:** Minimum inhibitory concentrations (MICs) were determined for *LpnSg1* clinical (n=100) and related environmental (n=7) isolates, collected between 2006-2022 in the context of the National Legionnaire’s Disease Surveillance Programme, against azithromycin, clarithromycin, erythromycin, levofloxacin, ciprofloxacin, moxifloxacin, rifampicin, doxycycline, tigecycline, and amoxicillin/clavulanic acid, using 3 different assays. Isolates were also PCR-screened for the presence of the *lpeAB* gene.

**Results:** Twelve isolates had azithromycin MICs above the EUCAST tentative highest WT MIC, 9 of which were *lpeAB* negative; for erythromycin and clarithromycin, all isolates tested within the susceptible range. The number of isolates with MICs above the tentative highest WT MIC for the remaining antibiotics was: ciprofloxacin: 7; levofloxacin: 17; moxifloxacin: 8; rifampicin: 11; doxycycline: 82; tigecycline: 4. EUCAST breakpoints are not available for amoxicillin/clavulanic acid. We estimated the ECOFFs and one isolate had a MIC 8-fold higher than the E-test ECOFF. Additionally, a clinical isolate generated three colonies growing on the E-test inhibition zone that resulted in MICs 4-fold higher than for the parental isolate.

**Conclusions:** We report, for the first time, elevated MICs against first-line and other antibiotics (including azithromycin, fluoroquinolones and amoxicillin/clavulanic acid commonly used to treat pneumonia patients in Portugal) in Portuguese *L. pneumophila* strains. Results point towards decreased susceptibility in circulating strains, justifying further investigation.

## Introduction

The gram-negative bacillus *Legionella pneumophila* is clinically associated with Legionnaires’ Disease (LD), a severe form of community-acquired pneumonia (CAP)^1^. If untreated, the case fatality rate of LD can be up to 10%^2^. *L. pneumophila* serogroup 1 (sg1) is responsible for the majority of cases worldwide, including in Europe^2^. Although LD is relatively sporadic in Europe, with high heterogeneity in reporting between countries, notification rates have been increasing from 1.4 to 2.2 cases/100000 population between 2016 and 2021^3^, and large outbreaks have also been reported in recent years^4–6^.

Since *Legionella* replicates intracellularly, the choice of therapeutics for LD is limited to antimicrobials which can penetrate cells such as macrolides or fluoroquinolones^7^. Azithromycin, levofloxacin, or moxifloxacin are recommended as first-line treatment of LD^8^, but β-Lactams such as amoxicillin are also frequently used as first option to treat patients with CAP^9^. In Portugal, the recommended therapeutics for previously healthy CAP patients include amoxicillin as first option, with azithromycin, clarithromycin, or doxycycline as alternatives^10^. In patients with comorbidities or with recent antibiotherapy, the recommendations are to administer amoxicillin in combination with azithromycin, clarithromycin, or doxycycline, and levofloxacin or moxifloxacin as alternatives^10^.

Although antibiotic resistance in *Legionella* has not yet been a subject for major concern, ciprofloxacin-resistant bacteria have been isolated from patients undergoing treatment^11,12^, and a reduced sensitivity to erythromycin and azithromycin has also been reported in clinical and environmental isolates^13–15^. In the case of azithromycin, the phenotype has been associated to point mutations in the *lpeAB* gene (coding for an efflux pump involved in macrolide resistance)^13,16^.

*In vitro* antibiotic susceptibility testing (AST) is crucial to determine the minimum inhibitory concentration (MIC) of the drugs and to assess whether bacteria are showing signs of resistance. However, there is no gold standard method recommended for *Legionella*. Available methods include agar dilution, broth microdilution (BMD) and gradient strip testing on buffered charcoal-containing yeast extract (BCYE) agar^7^. BMD is usually considered the most reliable method for clinically relevant bacteria, but it is time-consuming due to the slow growth rate and complex medium requirements of *Legionella*. Gradient strips are more widely used due to their ease of use, although they tend to produce MIC values that are higher than those returned by BMD in consequence of the use of charcoal in the medium. Recently a new method using a solid charcoal-free medium (LASARUS)^17^ has shown to produce results more in line with those of BMD, but it still needs further validation. The European Committee on Antimicrobial Susceptibility Testing (EUCAST) has produced guidelines for the interpretation of MICs in *Legionella* using either the BMD or the gradient strip methods^18^, based on the highest MIC observed in wild type (WT) isolates from published studies. These values are used as a threshold for submitting the isolates to a reference laboratory for further testing, but there is no universal agreement on epidemiological cut-off (ECOFF) values to discriminate between wild-type and potentially resistant strains partly due to the different MIC values returned by different *in vitro* methods^19^. Additionally, there are currently no clinical breakpoints available for *Legionella* to define whether an infection is likely to be treatable or not in a patient^20^. Given the increasing trend in LD notifications and its severity, there is a need for a more extensive antimicrobial susceptibility screening of both clinical and environmental *L. pneumophila* strains to have a clearer picture of the situation at the European level. Additionally, comparing different *in vitro* AST methods is pivotal to help refining and standardising the current methods and guidelines for the determination and interpretation of MICs and cut-off values for *Legionella*.

There is little information on the antibiotic susceptibility profile of *L. pneumophila* strains circulating in Portugal. A study from 1997 including both clinical and environmental isolates did not find evidence of reduced susceptibility^21^. However, a more recent study involving strains isolated from water samples found evidence of potential resistance to levofloxacin^22^.

Our study aimed to determine the antibiotic susceptibility of *L. pneumophila* serogroup 1 clinical and environmental isolates collected in Portugal between 2006 and February 2022 to ten antibiotics used in the clinical practice. Three AST methods were used (gradient strip, BMD, and LASARUS agar medium). Additionally, we determined the prevalence of the *lpeAB* gene.

## Materials and methods

### Bacterial strains

A total of 107 *L. pneumophila* sg1 isolates were tested: 100 clinical (of which 72 from sporadic LD cases, and 28 from 11 confirmed outbreaks) and seven environmental (associated with seven of the 11 confirmed outbreaks). The isolates were collected between 2006 and February 2022 in the context of the National Legionnaire’s Disease Surveillance Programme and stored at the National Reference Laboratory for *Legionella* of the National Institute of Health Doutor Ricardo Jorge (NRL/INSA) in Lisboa, Portugal. Briefly, isolates stored at <-70°C were inoculated on buffered charcoal-containing yeast extract medium supplemented with α-ketoglutarate (BCYE-α) and incubated at 36±2°C in a humid chamber for 48-72 h before testing. The fully susceptible *L. pneumophila* subsp. *pneumophila* Philadelphia-1 strain (ATCC 33152) was inoculated in the same way and used as a reference strain.

### Antimicrobial susceptibility testing

Bacteria were tested for the following ten antibiotics: azithromycin, clarithromycin, erythromycin, levofloxacin, ciprofloxacin, moxifloxacin, rifampicin, doxycycline, tigecycline, and amoxicillin/clavulanic acid (2/1). AST was performed by three methods. The gradient strip method was performed at the NRL/INSA following the recommendations of EUCAST^18^ and using E-test® gradient strips (bioMérieux SA, France) according to the manufacturer’ instructions^23^. All isolates were also shipped to the Department of Medical Microbiology, Cardiff University School of Medicine, United Kingdom, where they were tested for the same antibiotics using the BMD and LASARUS agar methods as previously described^17^. MICs were read as the lowest antimicrobial concentration inhibiting growth. Additionally, DNA was extracted from all the isolates and used for the PCR amplification of the *lpeAB* gene as previously described^13^.

### Data analysis

MIC values were used to classify the isolates as susceptible (MIC below or equal to the tentative highest WT MIC) or with reduced susceptibility (MIC above the tentative highest WT MIC) according to the EUCAST guidelines^18^. MIC values were used to calculate the minimum concentration at which 50% (MIC_50_) and 90% (MIC_90_) of the isolates are inhibited, respectively, and the MIC range. Additionally, ECOFFs (95%) were calculated using the ECOFFinder program (version 2.1)^24,25^. MIC and ECOFF (if available) values were extrapolated from other representative published studies for comparison. Data were tabulated and graphs were constructed using Microsoft Excel.

### Ethical approvals

Bacterial isolates obtained from anonymised sources arising from routine diagnostic samples were used. As no patient identifying information was available to the investigators, this project represents a service evaluation and development of future diagnostic tools; therefore, no ethical approval was required.

## Results

The MIC distribution for the tested antibiotics in the 107 isolates is shown in Table 1, while a summary of the number of isolates showing reduced susceptibility to the antibiotics is shown in Table 2. The MIC range, MIC_50_, MIC_90_ and ECOFF values are reported in Table 3. The latter table also reports the values from representative studies as a comparison. The mode of deviation of the MIC values obtained by gradient strip and LASARUS compared to the BMD gold standard are shown in Figure 1. All the raw MIC values and additional information about the isolates are available on Zenodo (DOI: 10.5281/zenodo.8367289). A total of nine (8.4%) isolates were found carrying the *lpeAB* gene.

**Table 1:**
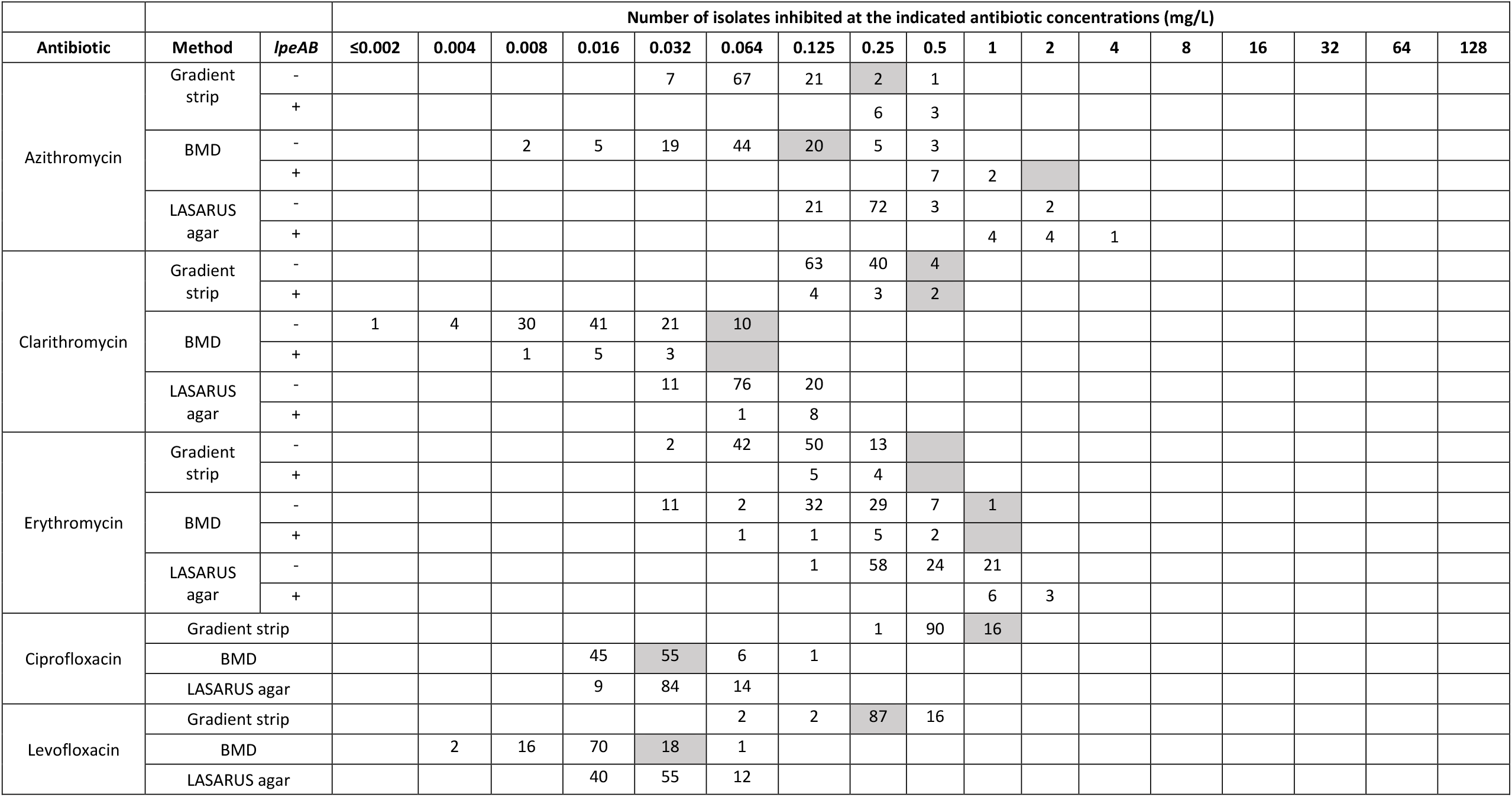
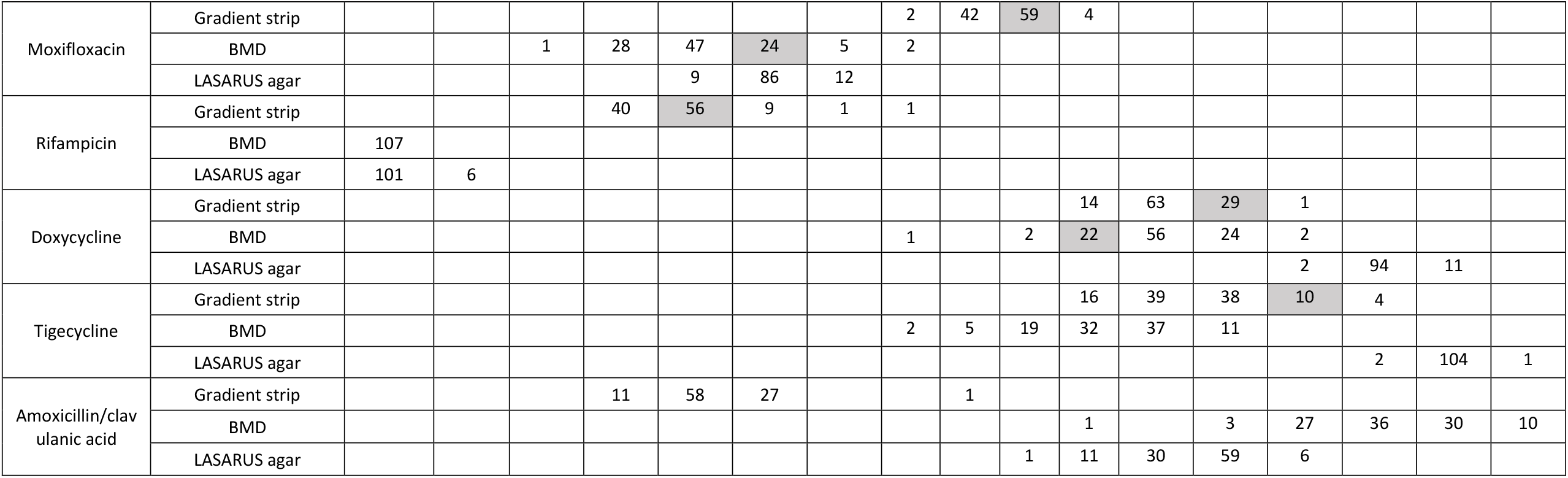
MIC distribution of the *L. pneumophila* serogroup 1 isolates from Portugal (n = 107). Tentative EUCAST highest WT MIC values are highlighted in grey.

|  |  |  | Number of isolates inhibited at the indicated antibiotic concentrations (mg/L) |  |  |  |  |  |  |  |  |  |  |  |  |  |  |  |  |
| --- | --- | --- | --- | --- | --- | --- | --- | --- | --- | --- | --- | --- | --- | --- | --- | --- | --- | --- | --- |
| Antibiotic | Method | *lpeAB* | ≤0.002 | 0.004 | 0.008 | 0.016 | 0.032 | 0.064 | 0.125 | 0.25 | 0.5 | 1 | 2 | 4 | 8 | 16 | 32 | 64 | 128 |
| Azithromycin | Gradient strip | - |  |  |  |  | 7 | 67 | 21 | 2 | 1 |  |  |  |  |  |  |  |  |
|  |  | + |  |  |  |  |  |  |  | 6 | 3 |  |  |  |  |  |  |  |  |
|  | BMD | - |  |  | 2 | 5 | 19 | 44 | 20 | 5 | 3 |  |  |  |  |  |  |  |  |
|  |  | + |  |  |  |  |  |  |  |  | 7 | 2 |  |  |  |  |  |  |  |
|  | LASARUS agar | - |  |  |  |  |  |  | 21 | 72 | 3 |  | 2 |  |  |  |  |  |  |
|  |  | + |  |  |  |  |  |  |  |  |  | 4 | 4 | 1 |  |  |  |  |  |
| Clarithromycin | Gradient strip | - |  |  |  |  |  |  | 63 | 40 | 4 |  |  |  |  |  |  |  |  |
|  |  | + |  |  |  |  |  |  | 4 | 3 | 2 |  |  |  |  |  |  |  |  |
|  | BMD | - | 1 | 4 | 30 | 41 | 21 | 10 |  |  |  |  |  |  |  |  |  |  |  |
|  |  | + |  |  | 1 | 5 | 3 |  |  |  |  |  |  |  |  |  |  |  |  |
|  | LASARUS agar | - |  |  |  |  | 11 | 76 | 20 |  |  |  |  |  |  |  |  |  |  |
|  |  | + |  |  |  |  |  | 1 | 8 |  |  |  |  |  |  |  |  |  |  |
| Erythromycin | Gradient strip | - |  |  |  |  | 2 | 42 | 50 | 13 |  |  |  |  |  |  |  |  |  |
|  |  | + |  |  |  |  |  |  | 5 | 4 |  |  |  |  |  |  |  |  |  |
|  | BMD | - |  |  |  |  | 11 | 2 | 32 | 29 | 7 | 1 |  |  |  |  |  |  |  |
|  |  | + |  |  |  |  |  | 1 | 1 | 5 | 2 |  |  |  |  |  |  |  |  |
|  | LASARUS agar | - |  |  |  |  |  |  | 1 | 58 | 24 | 21 |  |  |  |  |  |  |  |
|  |  | + |  |  |  |  |  |  |  |  |  | 6 | 3 |  |  |  |  |  |  |
| Ciprofloxacin | Gradient strip |  |  |  |  |  |  |  |  | 1 | 90 | 16 |  |  |  |  |  |  |  |
|  | BMD |  |  |  |  | 45 | 55 | 6 | 1 |  |  |  |  |  |  |  |  |  |  |
|  | LASARUS agar |  |  |  |  | 9 | 84 | 14 |  |  |  |  |  |  |  |  |  |  |  |
| Levofloxacin | Gradient strip |  |  |  |  |  |  | 2 | 2 | 87 | 16 |  |  |  |  |  |  |  |  |
|  | BMD |  |  | 2 | 16 | 70 | 18 | 1 |  |  |  |  |  |  |  |  |  |  |  |
|  | LASARUS agar |  |  |  |  | 40 | 55 | 12 |  |  |  |  |  |  |  |  |  |  |  |

|  |  |  |  |  |  |  |  |  |  |  |  |  |  |  |  |  |  |  |
| --- | --- | --- | --- | --- | --- | --- | --- | --- | --- | --- | --- | --- | --- | --- | --- | --- | --- | --- |
| Moxifloxacin | Gradient strip |  |  |  |  |  |  |  | 2 | 42 | 59 | 4 |  |  |  |  |  |  |
|  | BMD |  |  | 1 | 28 | 47 | 24 | 5 | 2 |  |  |  |  |  |  |  |  |  |
|  | LASARUS agar |  |  |  |  | 9 | 86 | 12 |  |  |  |  |  |  |  |  |  |  |
| Rifampicin | Gradient strip |  |  |  | 40 | 56 | 9 | 1 | 1 |  |  |  |  |  |  |  |  |  |
|  | BMD | 107 |  |  |  |  |  |  |  |  |  |  |  |  |  |  |  |  |
|  | LASARUS agar | 101 | 6 |  |  |  |  |  |  |  |  |  |  |  |  |  |  |  |
| Doxycycline | Gradient strip |  |  |  |  |  |  |  |  |  |  | 14 | 63 | 29 | 1 |  |  |  |
|  | BMD |  |  |  |  |  |  |  | 1 |  | 2 | 22 | 56 | 24 | 2 |  |  |  |
|  | LASARUS agar |  |  |  |  |  |  |  |  |  |  |  |  |  | 2 | 94 | 11 |  |
| Tigecycline | Gradient strip |  |  |  |  |  |  |  |  |  |  | 16 | 39 | 38 | 10 | 4 |  |  |
|  | BMD |  |  |  |  |  |  |  | 2 | 5 | 19 | 32 | 37 | 11 |  |  |  |  |
|  | LASARUS agar |  |  |  |  |  |  |  |  |  |  |  |  |  |  | 2 | 104 | 1 |
| Amoxicillin/clavulanic acid | Gradient strip |  |  |  | 11 | 58 | 27 |  |  | 1 |  |  |  |  |  |  |  |  |
|  | BMD |  |  |  |  |  |  |  |  |  |  | 1 |  | 3 | 27 | 36 | 30 | 10 |
|  | LASARUS agar |  |  |  |  |  |  |  |  |  | 1 | 11 | 30 | 59 | 6 |  |  |  |

**Table 2:** Summary of the number and percentage of isolates with MIC above the EUCAST highest WT MIC in the *L. pneumophila* serogroup 1 isolates from Portugal (n = 107)

|  | No. isolates with MIC (mg/L) > EUCAST highest WT MIC (%) |  |  |  |
| --- | --- | --- | --- | --- |
| Antibiotic | By gradient strip | By BMD | By both | Total |
| Azithromycin | 4 (3.7) | 8 (7.5) | 0 (0) | **12 (11.2)** |
| Ciprofloxacin | 0 (0) | 7 (6.5) | 0 (0) | **7 (6.5)** |
| Levofloxacin | 16 (14.9) | 1 (0.9) | 0 (0) | **17 (15.9)** |
| Moxifloxacin | 1 (0.9) | 4 (3.7) | 3 (2.8) | **8 (7.5)** |
| Rifampicin | 11 (10.3) | 0 (0) | 0 (0) | **11 (10.3)** |
| Doxycycline | 0 (0) | 81 (75.7) | 1 (0.9) | **82 (76.6)** |
| Tigecycline | 4 (3.7) | 0 (0) | 0 (0) | **4 (3.7)** |

**Table 3:**
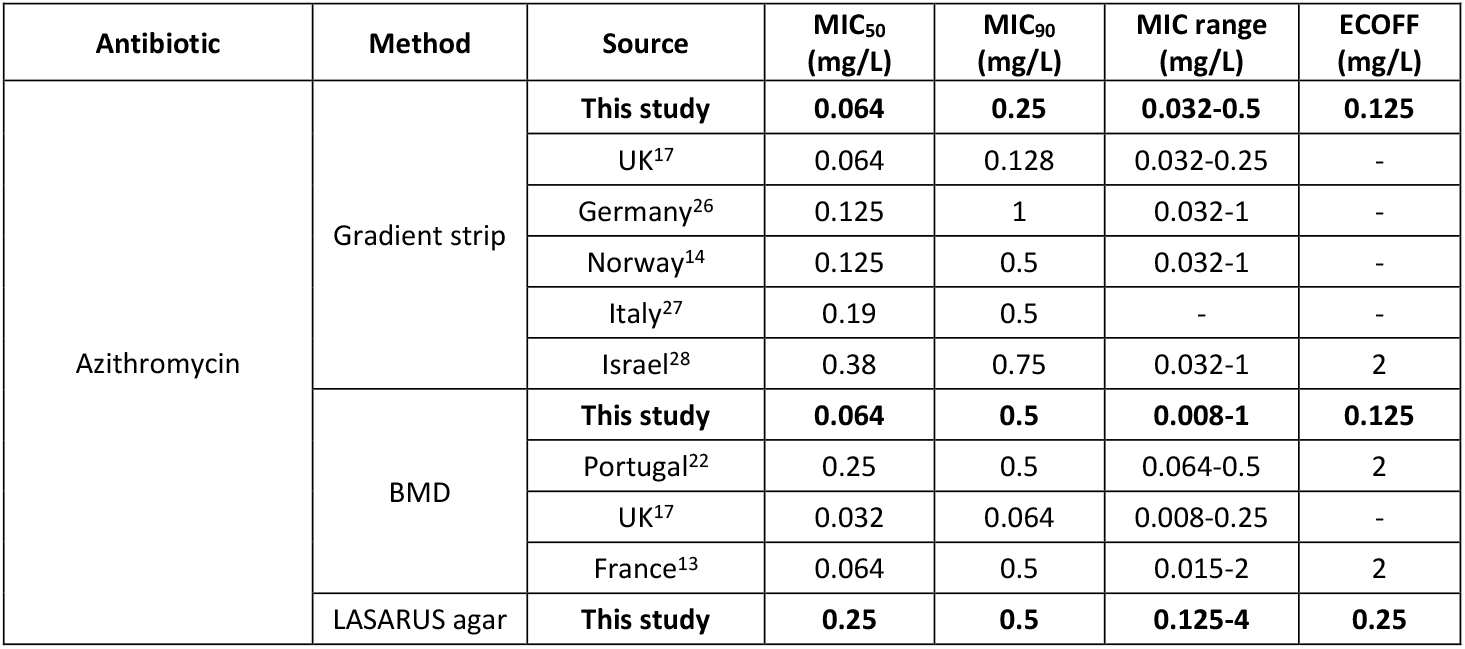
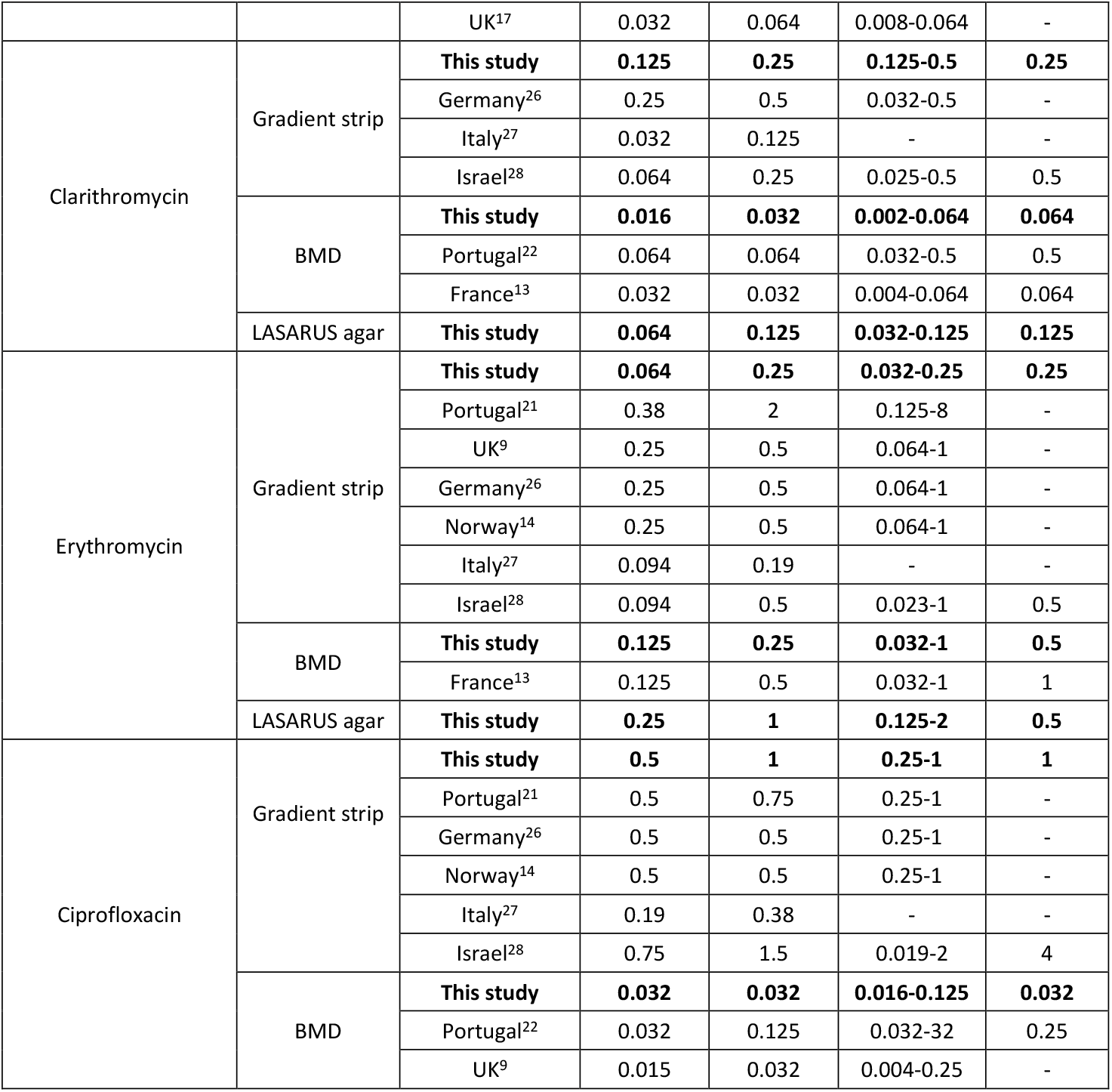
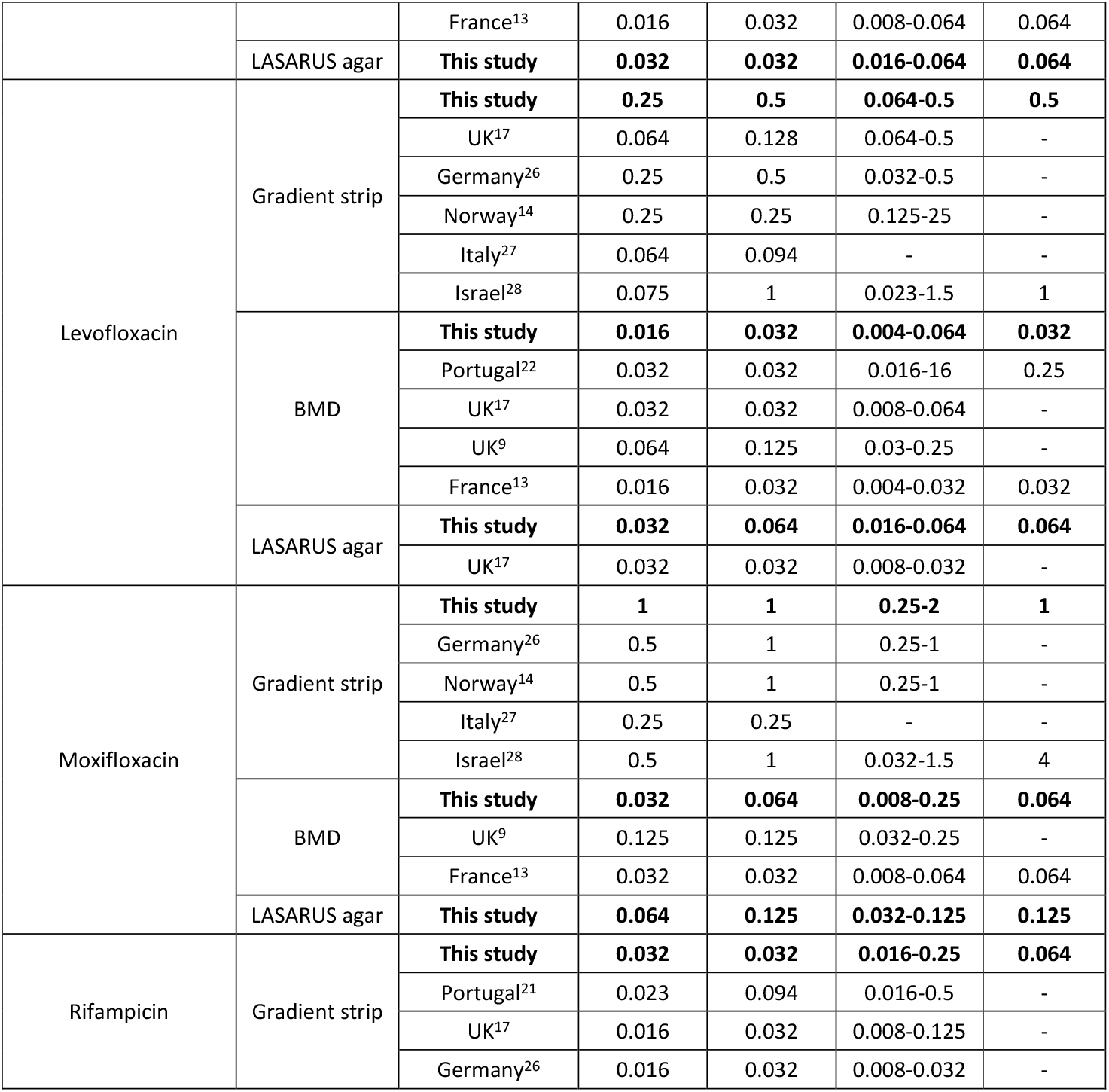
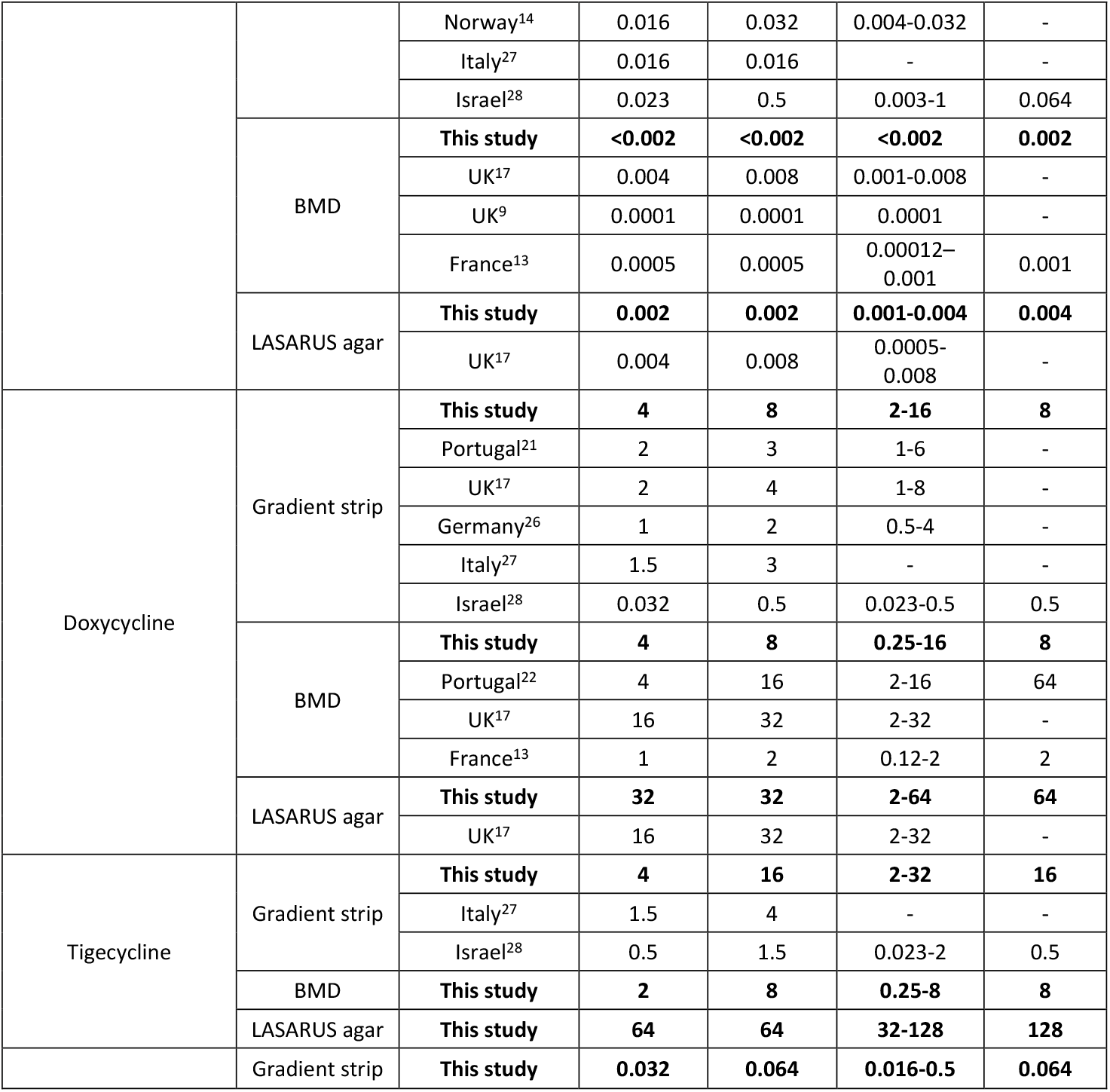
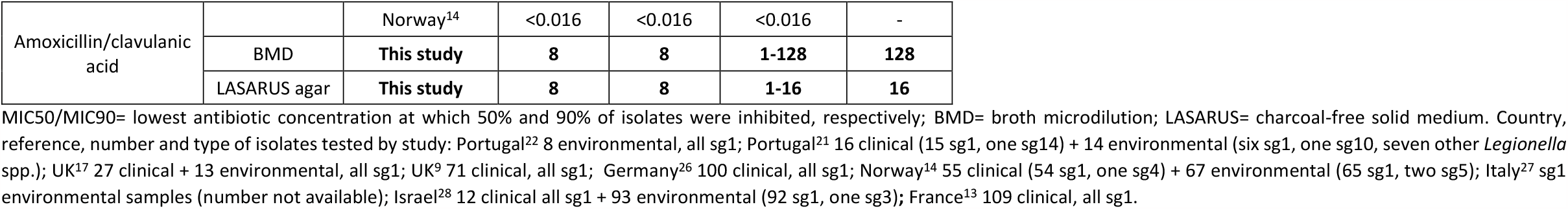
Minimum inhibitory concentration ranges, MIC_50_, MIC_90_ and ECOFF values of the *L. pneumophila* serogroup 1 isolates from Portugal (n = 107) compared to other representative studies.

**Figure 1:**
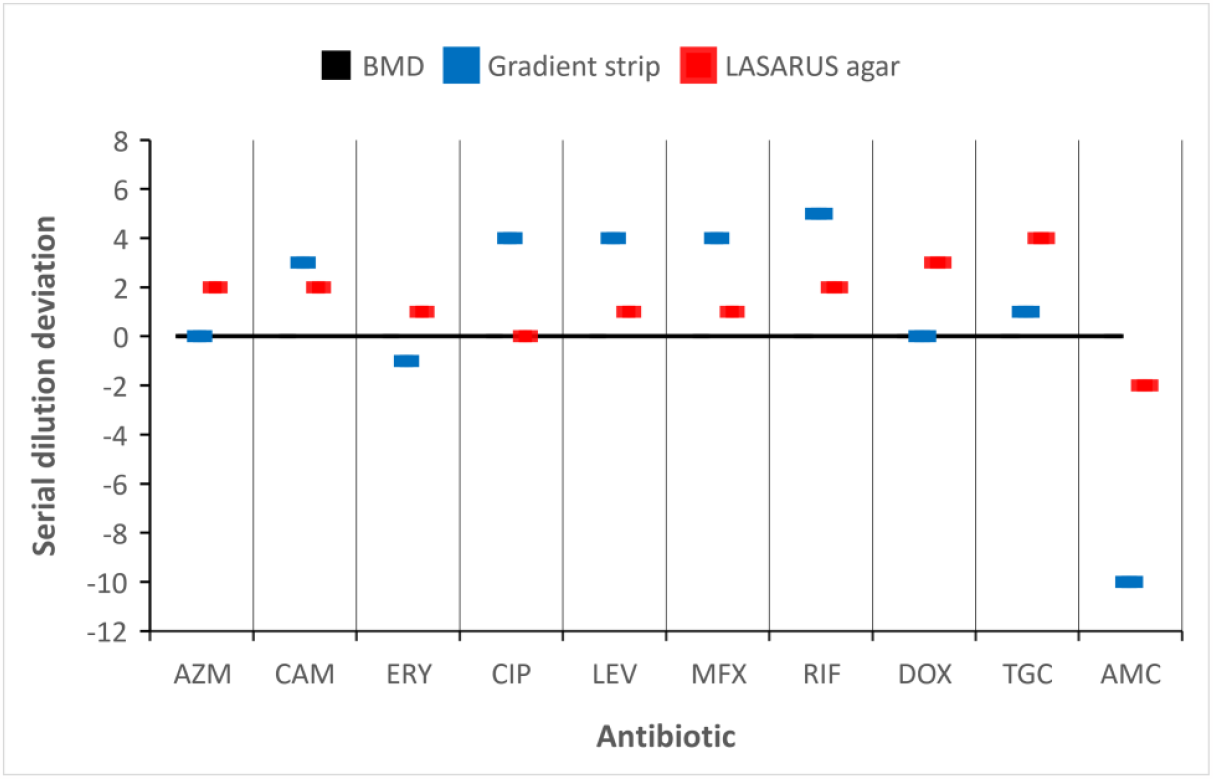
Modal averages of MIC values by gradient strip and LASARUS agar expressed as serial dilution deviation from the BMD gold standard (black line). AZM= azithromycin; CAM= clarithromycin; ERY= erythromycin; CIP= ciprofloxacin; LEV= levofloxacin; MFX= moxifloxacin; RIF= rifampicin; DOX= doxycycline; TGC= tigecycline; AMC= amoxicillin/clavulanic acid.

### Macrolides

In total, 12 isolates (11.2%) had azithromycin MICs above the tentative highest WT MIC: four (3.7%) by gradient strip (one *lpeAB*-negative and three *lpeAB*-positive) and eight (7.5%) by BMD (threshold 0.125 mg/L, all *lpeAB*-negative). For erythromycin and clarithromycin, all isolates tested within the susceptible range by both the gradient test and BMD. Estimated ECOFF values using ECOFF finder in our study were two-fold lower than the highest WT MIC values published by EUCAST^18^ with the gradient strip for all three macrolides and with the BMD only for erythromycin.

### Fluoroquinolones

Isolates with MICs above the tentative highest WT MIC for the tested fluoroquinolones were found: seven (6.5%) for ciprofloxacin (all BMD, MIC >0.032 mg/L), 17 (15.9%) for levofloxacin (16 for gradient strip MIC >0.25; one for BMD MIC >0.032 mg/L), and eleven (7.5%) for moxifloxacin (four for BMD MIC >1; seven for BMD MIC >0.064). By considering either gradient strip or BMD results, a total of four (3.7%) isolates had reduced susceptibility for all three fluoroquinolones and an additional three (2.8%) had reduced susceptibility for both ciprofloxacin and moxifloxacin. The ECOFF value for levofloxacin using ECOFF finder obtained with gradient strip was two-fold higher than the highest WT MIC values published by EUCAST^18^, while the BMD ECOFFs corresponded to putative susceptibility thresholds suggested by EUCAST.

### Rifampicin

A total of 11 (10.3%) isolates had a MIC above the tentative highest WT MIC for rifampicin (only for by gradient strip MIC >0.032 mg/L). Our BMD MICs corresponded to values indicated by EUCAST^18^, while the ECOFF determined by ECOFF finder for the gradient strip was two-fold higher than EUCAST recommends.

### Tetracyclines

Overall, 82 isolates (76.6%) had doxycycline MICs above the tentative highest WT MIC (one for gradient strip MIC >8 mg/L, the rest were BMD MIC >2 mg/L), and four of these (3.7% out of the total tested) also had reduced susceptibility for tigecycline (gradient strip MIC >16; no value available for BMD to assess). The ECOFF from ECOFF finder for both antibiotics for gradient strip corresponded to highest WT MIC from EUCAST, while the ECOFF for doxycycline for BMD was four-fold higher than identified by EUCAST.

### Amoxicillin/clavulanic acid

Tentative EUCAST tentative highest WT MIC values for this antibiotic are not available for *Legionella*, but considering the gradient strip test and the relative estimated ECOFF (0.064 mg/L), one isolate (0.9%) had a MIC above it. During testing of one of the clinical isolates (E206) with the gradient strip test, three colonies were observed growing within the inhibition halo. The colonies were then re-isolated and tested separately. All three colonies returned a MIC two-to four-fold higher than that observed for the initial isolate by both the gradient strip and BMD tests, and 16-fold higher with the LASARUS agar test. The three colonies also showed elevated MICs compared to the initial isolate for the following antibiotics: ciprofloxacin (four-fold higher by both BMD and LASARUS agar), levofloxacin (two-fold higher in one colony by BMD and in all three by LASARUS agar; four-fold higher in two colonies by BMD only), and both azithromycin and clarithromycin (two-fold higher by BMD only). The three colonies also showed a two-fold lower MIC for tigecycline compared to the initial isolate (by BMD only), and one colony had a two-fold lower MIC for azithromycin.

### Reduced susceptibility to multiple antibiotics

By taking into account either gradient strip of BMD results, 16 isolates (14.9%) had reduced susceptibility to both levofloxacin and doxycycline, 12 (11.2%) to azithromycin and doxycycline, and 10 (9.3%) to rifampicin and doxycycline. Other occurrences included reduced susceptibility to ciprofloxacin/moxifloxacin and doxycycline (six isolates each, 5.6%), levofloxacin and rifampicin/tigecycline (four isolates each, 3.7%), rifampicin and tigecycline (two isolates, 1.9%), and azithromycin and rifampicin (one isolate, 0.9%).

### Comparison of MICs between the three AST methods

Compared to the BMD gold standard (Figure 1), the gradient strip returned significantly elevated thresholds of inhibition for clarithromycin (three serial dilutions), all fluoroquinolones (four dilutions), and rifampicin (five dilutions). The gradient strip also showed a significantly reduced threshold (down to 10 dilutions) compared to BMD for amoxicillin/clavulanic acid. The LASARUS agar method returned MIC values which were more comparable to those by BMD, with all thresholds of inhibition showing a deviation of plus/minus two dilution factors except for tigecycline (four dilutions). The deviations of gradient strip and LASARUS results from the BMD are reflected also by MIC ranges, MIC_50_, MIC_90_ and ECOFF values (Table 2).

## Discussion

Our study generated the first data on the antibiotic susceptibility profile of *L. pneumophila* serogroup 1 isolated from Portuguese LD patients since 1997^21^. Compared to a recent Portuguese study using only broth microdilution^22^, we used in addition another method recommended by EUCAST (gradient strip) and the recently proposed described LASARUS agar dilution method ^17^.

Overall, the MIC values and ranges of our isolates were comparable (with minor variations) to those reported in other studies using the gradient strip and/or the BMD methods. Rifampicin was the most effective antibiotic, while doxycycline and tigecycline were the least effective. These results are in accordance to those reported by other authors^13,17,26,27,29^. To classify the isolates as susceptible or (potentially) resistant we used the MIC thresholds recommended by EUCAST for referring isolates to reference laboratories as putatively resistant^18^. It is important to note that these reference values are based on literature review and differ between the gradient strip and BMD approaches. While an isolate can have exactly the same MIC by both methods for some antibiotics, it can be differentially classified as susceptible because the literature for one method shows a higher average range. Nevertheless, we found evidence of reduced susceptibility to various antibiotics, including to first-line compounds.

We found 12 isolates with reduced susceptibility to azithromycin. Interestingly, nine of these isolates were not carrying the *lpeAB* gene which is known to confer resistance to macrolides^13,14,16^, and we did not have any information regarding treatment of the patient. As expected most of (but not all) the isolates carrying this gene had the highest MICs. The lack of this gene in isolates with high azithromycin MIC is not unexpected and it has been previously reported^17,30^, including in Portuguese environmental isolates^22^, suggesting that other resistance/reduced susceptibility mechanisms might be involved. Mutants of *L. pneumophila* sg1 selected *in vitro* for resistance to macrolides showed MIC values above 16 mg/L by BMD^13,18^. Although none of our isolates had MIC values comparable to these, since azithromycin is expected to be one of the most frequent antibiotic administered to LD patients following standardised treatment guidelines, it will be important to continue to assess and monitor potential azithromycin resistance phenomena in Portugal. Although isolates did not show any reduced susceptibility for the other two macrolides tested, all the isolates with the *lpeAB* gene had MICs at the higher end of the range for erythromycin.

Some isolates also showed reduced susceptibility to fluoroquinolones, including four with MIC values above the breakpoints for all the antibiotics tested. Two isolates from our sample had a BMD MIC of 0.250 mg/L for moxifloxacin. Previous studies have reported a ciprofloxacin-resistant *L. pneumophila* strain isolated from a patient, showing a MIC of 2 mg/L by gradient strip^11^, and *in vitro* selected strains showing MIC values above 0.125 mg/L by BMD for either levofloxacin or moxifloxacin^13,18^.

We found 11 isolates with reduced susceptibility to rifampicin (including two with a MIC four- and eight-fold higher than the ECOFF, respectively) by the gradient strip method. Using the same approach, a previous study reported isolates with MIC values up to 4 mg/L^27^.

The majority (more than 70%) of our isolates showed very high inhibition thresholds for doxycycline by BMD (above the EUCAST reference thresholds for submission to reference laboratories as putatively resistant), although similar MIC values have been reported in UK^17^ and Portuguese^22^ environmental isolates before. Whether these values reflect naturally occurring variation in susceptibility, or the presence of resistance, requires further testing and analysis.

One of the most interesting results of our study came from the amoxicillin/clavulanic acid testing. No breakpoints are available for this antibiotic combination in *Legionella*, and to the best of our knowledge only one study assessed the susceptibility of these bacteria to it^14^. Compared to the gradient strip results from that study (all isolates had a MIC <0.016 mg/L)^14^, 80% of our isolates had MIC values or 0.032 mg/L and above. We also managed to isolate three colonies showing signs of resistance to amoxicillin/clavulanic acid, as confirmed by a significantly increased threshold of inhibition compared to the parent isolate, not only by gradient strip but also by BMD. These three isolates underwent whole-genome sequencing and were confirmed to be the same as the parental isolate (National Reference Laboratory for *Legionella, pers. comm*.). Further analysis is underway to determine the potential molecular mechanism behind the phenotype. The finding that *L. pneumophila* can develop resistance to this antibiotic is very relevant for patient management and public health. Beta-lactam antibiotics such as amoxicillin can be frequently used to treat patients with CAP^9^, and this drug was the second most frequently reported as treatment for CAP patients according to surveillance data collected non-systematically at the National Reference Laboratory relative to the period of this study. However, it is important to note that beta-lactams would only be effective against extracellular Legionella and are unlikely to eradicate infection on its own.

We also estimated the epidemiological cut-off (ECOFF) values specifically in our sample to compare them to the EUCAST tentative highest WT MICs. While the ECOFFs obtained with BMD largely overlapped with the EUCAST threshold values (apart from being lower for erythromycin, and higher for doxycycline), more discrepancies were observed for values obtained with the gradient strips. While values overlapped for tetracyclines and fluoroquinolones (except levofloxacin), ECOFFs were lower than the ECOFFs for all macrolides and, conversely, higher for levofloxacin and rifampicin. Discrepancies have been reported in another study from Portugal^22^. Universal ECOFF values have not been formally assigned making difficult to ascertain wild-type and resistant field strains, and more data are needed for reaching a much-needed international standardization^7^.

While we observed relatively concordant MIC results between BMD and LASARUS for fluoroquinolones, rifampicin, and amoxicillin/clavulanic acid, the BMD results were more concordant with the gradient strip results for macrolides (with the exception of clarithromycin) and tetracyclines. Compared to the other two methods and similarly to a previous study^17^, the gradient strip returned more elevated MIC values for both fluoroquinolones and rifampicin. This result can be partially explained by the known chelating effect of activated charcoal in the BCYE medium used in the Gradient strips. The degree of antimicrobial compound adsorption in a charcoal-containing medium can increase the MIC values, as reported by various studies^31–33^. This phenomenon is not expected in the LASARUS. However, we also reported significantly lower MIC values for amoxicillin/clavulanic acid using the gradient strip compared to the other two methods. Differences between antibiotics in the degree of their absorption and bioavailability in different media cannot be excluded. In order to surpass the known constraints of the gradient strip and the time-consuming and logistically difficult BMD approaches, the LASARUS medium offers some interesting advantages^17^: it is charcoal-free, and it allows inoculation of multiple samples using a multipoint inoculator. Additionally, it is a translucent medium allowing an easier and safer reading of the results (which could also be automated using optical readers). The gradient strips are not compatible with the current formulation of the LASARUS medium^17^, so a combination of the two approaches is currently not possible. As shown already by another study^17^, and confirmed by our data, overall the LASARUS medium looks as a promising alternative to BMD.

This study had some limitations. Only fraction of the total archived isolates at the National Reference Laboratory were tested. We tested mostly clinical samples from LD patients, so the resulting picture may not be fully representative of *L. pneumophila* bacteria circulating in Portugal (particularly in the environment). Information about antibiotic treatment was available for only around 20% of the isolates, and we cannot infer on the impact of potential antibiotic treatments administered to patients on the results of our assays. However, for the isolates for which the information was available, we did not observe any correlation between the type of treatment and the presence of reduced susceptibility to the corresponding compound.

Our results highlight the need for more extensive AST data on *L. pneumophila* in Portugal. Given the severity and potentially fatal outcome of LD, it is important to monitor the antimicrobial susceptibility status of circulating bacterial strains to identify the emergence of resistance in a timely manner. Whole genome sequencing of isolates with reduced antibiotic susceptibility will also be of great help to elucidate potential molecular determinants affecting the phenotype and giving rise to resistant strains.

## Data Availability

All data produced are available online at Zenodo (DOI: 10.5281/zenodo.8367289)

## Acknowledgements

The authors would like to acknowledge and express their gratitude to the General Directorate of Health and local health authorities, and all clinicians and pathologists who have contributed to the laboratory component of the National Legionnaire’s Disease Surveillance Programme. Corrado Minetti is a fellow of the ECDC Fellowship Programme, supported financially by the European Centre for Disease Prevention and Control. The views and opinions expressed herein do not state or reflect those of ECDC. ECDC is not responsible for the data and information collation and analysis and cannot be held liable for conclusions or opinions drawn.

## References

1. Cunha BA, Burillo A, Bouza E. Legionnaires’ disease. The Lancet 2016; 387: 376–85.

2. Beauté J, Plachouras D, Sandin S, Giesecke J, Sparén P. Healthcare-Associated Legionnaires’ Disease, Europe, 2008-2017. Emerg Infect Dis 2020; 26: 2309–18.

3. European Centre for Disease Prevention and Control. Legionnairés Disease [Internet]. Surveillance Atlas of Infectious Diseases. Available at: https://atlas.ecdc.europa.eu/public/index.aspx. Accessed July 21, 2022.

4. Shivaji T, Pinto CS, San-Bento A, et al. A large community outbreak of Legionnaires’ disease in Vila Franca de Xira, Portugal, October to November 2014. Eurosurveillance 2014; 19: 20991.

5. Faccini M, Russo AG, Bonini M, et al. Large community-acquired Legionnaires’ disease outbreak caused by Legionella pneumophila serogroup 1, Italy, July to August 2018. Euro Surveill Bull Eur Sur Mal Transm Eur Commun Dis Bull 2020; 25.

6. Maisa A, Brockmann A, Renken F, et al. Epidemiological investigation and case–control study: a Legionnaires’ disease outbreak associated with cooling towers in Warstein, Germany, August–September 2013. Eurosurveillance 2015; 20: 30064.

7. Portal E, Descours G, Ginevra C, et al. Legionella antibiotic susceptibility testing: is it time for international standardization and evidence-based guidance? J Antimicrob Chemother 2021; 76: 1113–6.

8. Viasus D, Gaia V, Manzur-Barbur C, Carratalà J. Legionnaires’ Disease: Update on Diagnosis and Treatment. Infect Dis Ther 2022; 11: 973–86.

9. Wilson RE, Hill RLR, Chalker VJ, Mentasti M, Ready D. Antibiotic susceptibility of Legionella pneumophila strains isolated in England and Wales 2007-17. J Antimicrob Chemother 2018; 73: 2757–61.

10. Direção-Geral da Saúde. Norma 045/2011 “Antibioterapia na Pneumonia Adquirida na Comunidade em Adultos Imunocompetentes”. 2011. Available at: https://www.dgs.pt/directrizes-da-dgs/normas-e-circulares-normativas/norma-n-0452011-de-26122011-jpg.aspx.

11. Bruin JP, Koshkolda T, IJzerman EPF, et al. Isolation of ciprofloxacin-resistant Legionella pneumophila in a patient with severe pneumonia. J Antimicrob Chemother 2014; 69: 2869–71.

12. Shadoud L, Almahmoud I, Jarraud S, et al. Hidden Selection of Bacterial Resistance to Fluoroquinolones In Vivo: The Case of Legionella pneumophila and Humans. EBioMedicine 2015; 2: 1179–85.

13. Vandewalle-Capo M, Massip C, Descours G, et al. Minimum inhibitory concentration (MIC) distribution among wild-type strains of Legionella pneumophila identifies a subpopulation with reduced susceptibility to macrolides owing to efflux pump genes. Int J Antimicrob Agents 2017; 50: 684–9.

14. Natås OB, Brekken AL, Bernhoff E, Hetland MAK, Löhr IH, Lindemann PC. Susceptibility of Legionella pneumophila to antimicrobial agents and the presence of the efflux pump LpeAB. J Antimicrob Chemother 2019; 74: 1545–50.

15. Ginevra C, Beraud L, Pionnier I, et al. Detection of highly macrolide-resistant Legionella pneumophila strains from a hotel water network using systematic whole-genome sequencing. J Antimicrob Chemother 2022; 77: 2167–70.

16. Massip C, Descours G, Ginevra C, Doublet P, Jarraud S, Gilbert C. Macrolide resistance in Legionella pneumophila: the role of LpeAB efflux pump. J Antimicrob Chemother 2017; 72: 1327–33.

17. Portal E, Sands K, Portnojs A, Chalker VJ, Spiller OB. Legionella antimicrobial sensitivity testing: comparison of microbroth dilution with BCYE and LASARUS solid media. J Antimicrob Chemother 2021; 76: 1197–204.

18. European Committee on Antimicrobial Susceptibility Testing. Guidance document on Antimicrobial Susceptibility Testing of Legionella pneumophila. May 2021. 2021. Available at: https://www.eucast.org/fileadmin/src/media/PDFs/EUCAST_files/Guidance_documents/Legionella_guidance_note_-_20210528.pdf.

19. Bruin JP, Ijzerman EPF, den Boer JW, Mouton JW, Diederen BMW. Wild-type MIC distribution and epidemiological cut-off values in clinical Legionella pneumophila serogroup 1 isolates. Diagn Microbiol Infect Dis 2012; 72: 103–8.

20. European Committee on Antimicrobial Susceptibility Testing. Breakpoint tables for interpretation of MICs and zone diameters. Version 13.0. 2023. Available at: https://www.eucast.org/fileadmin/src/media/PDFs/EUCAST_files/Breakpoint_tables/v_13.0_Breakpoint_Tables.pdf.

21. Marques T, Piedade J. Susceptibility testing by E-test and agar dilution of 30 strains of Legionella spp. isolated in Portugal. Clin Microbiol Infect 1997; 3: 365–8.

22. Cruz C, Rodrigues L, Fernandes F, Santos R, Paixão P, Chasqueira MJ. Antibiotic susceptibility pattern of Portuguese environmental Legionella isolates. Front Cell Infect Microbiol 2023; 13: 1141115.

23. Anon. ETEST® Package Insert. Available at: https://www.ilexmedical.com/files/E-test-Package-Insert/AST_WW.pdf.

24. European Committee on Antimicrobial Susceptibility Testing. MIC and zone distributions and ECOFFs. 2023. Available at: https://www.eucast.org/mic_and_zone_distributions_and_ecoffs. Accessed May 30, 2023.

25. Turnidge J, Kahlmeter G, Kronvall G. Statistical characterisation of bacterial wild-type MIC value distributions and the determination of epidemiological cut-off values. Clin Microbiol Infect Off Publ Eur Soc Clin Microbiol Infect Dis 2006; 12: 418–25.

26. Koshkolda T, Lück C. Antibiotic susceptibility of clinical Legionella pneumophila serogroup 1 strains isolated in Germany. J Antimicrob Chemother 2018; 73: 541–2.

27. De Giglio O, Napoli C, Lovero G, et al. Antibiotic susceptibility of Legionella pneumophila strains isolated from hospital water systems in Southern Italy. Environ Res 2015; 142: 586–90.

28. Sharaby Y, Nitzan O, Brettar I, Höfle MG, Peretz A, Halpern M. Antimicrobial agent susceptibilities of Legionella pneumophila MLVA-8 genotypes. Sci Rep 2019; 9: 6138.

29. Pappa O, Chochlakis D, Sandalakis V, Dioli C, Psaroulaki A, Mavridou A. Antibiotic Resistance of Legionella pneumophila in Clinical and Water Isolates—A Systematic Review. Int J Environ Res Public Health 2020; 17: 5809.

30. Cocuzza CE, Martinelli M, Perdoni F, et al. Antibiotic Susceptibility of Environmental Legionella pneumophila Strains Isolated in Northern Italy. Int J Environ Res Public Health 2021; 18: 9352.

31. Ruckdeschel G, Dalhoff A. The in-vitro activity of moxifloxacin against Legionella species and the effects of medium on susceptibility test results. J Antimicrob Chemother 1999; 43 Suppl B: 25–9.

32. García MT, Pelaz C, Giménez MJ, Aguilar L. In vitro activities of gemifloxacin versus five quinolones and two macrolides against 271 Spanish isolates of Legionella pneumophila: influence of charcoal on susceptibility test results. Antimicrob Agents Chemother 2000; 44: 2176–8.

33. Nielsen K, Bangsborg JM, Høiby N. Susceptibility of Legionella species to five antibiotics and development of resistance by exposure to erythromycin, ciprofloxacin, and rifampicin. Diagn Microbiol Infect Dis 2000; 36: 43–8.

